# Suicide after cancer diagnosis among older adults: A nationwide study from Austria

**DOI:** 10.64898/2026.07.14.26358049

**Authors:** Erwin Stolz, Anna Schultz, Emilise Lucerne Pötz, Maria Anna Smolle, Carlos Watzka, Christian Jagsch, Thomas Niederkrotenthaler, Annette Erlangsen

## Abstract

**Background:** Onset of cancer is linked to psychological distress and cancer is prevalent in older adults. Yet, the association to suicide is scarcely examined. The aim of this study was to assess whether cancer diagnosed in older adults is associated with suicide incidence.

**Methods:** All older adults (65+ years) who lived in Austria in the years 2014-2021 (n=2,175,134) were followed. Of these, 223,932 were diagnosed with a new cancer. We used non-parametric survival models with inverse-probability-treatment weights to compare risk ratios (relative risk) and risk differences (absolute risk) of older adults with and without cancer.

**Results:** Out of 2,158 suicide deaths, 442 (20.5%; 83.7% males) occurred among older adults with a new cancer diagnosis. The incidence rate was 74 among those with a new cancer diagnosis versus 23 per 100,000 person-years among those with no new cancer. One year after being diagnosed, older adults with a new cancer had a 4 times higher relative risk of dying by suicide compared to those without. The risk was highest within the first three months after diagnosis and for cancers with a poor prognosis (disseminated disease; lung, oesophagus, stomach, liver, pancreas, and brain cancers). The absolute risk of dying by suicide within 5 years after cancer diagnosis was 0.18% versus to 0.11% among those with no new cancer.

**Discussion:** Older adults who received a new cancer diagnosis had elevated suicide risks. Provision of support to cope with mental distress should be considered at cancer diagnosis, especially for older adults with a poor prognosis.

**HIGHLIGHTS:**

- 1 year after cancer diagnosis, suicide risk was 4-times higher than in the general population
- Suicide risk was highest in the first three months after cancer diagnosis but remained elevated throughout 5 years
- Suicide risk was higher for cancers with a poor prognosis
- Provision of psychosocial support should be considered immediately after diagnosis

## INTRODUCTION

Suicide rates among older adults are in many countries the highest of all age groups^1^, and within Europe, Austria has one of the highest old-age suicide rates.^2^ Cancers are relatively frequent among older adults^3^, and being diagnosed with cancer is associated with psychological distress^4^, and has been linked to elevated risk of suicide^5^. As summarized in a recent umbrella review^6^, individuals with cancer have an overall 50-70% higher risk of dying by suicide when compared to the general population. Excess risks of suicide have been shown for the first year after cancer diagnosis, specific cancer sites (e.g., oesophagus, lung, pancreas, stomach), cases with a poor prognosis, and among males, and older adults.^5,6^

Being diagnosed with cancer has been highlighted as a particular risk factor for suicide among older adults. Based on national data from Denmark and out of 39 physical diseases, high risks of suicide among older adults were found for lung cancer, gastrointestinal cancer, and bladder cancer.^7^ Further, suicide rates among individuals with cancers have been found to increase relative with increasing age; while those with cancer aged below 40 years had a suicide rate of 19 per 100,000, the rate for those aged 80 years and older was 44 per 100,000.^5^ A recent Danish study^8^ found that risk of suicide death after cancer diagnosis was 11-times higher among older adults aged 70+ years compared to those aged 18-39 years. Among somatic conditions, cancer was the only significant predictors of suicide among older adults 65+ in adjusted analyses of US data; having an effect size on par with affective and personality disorders.^9^ Similarly, evidence from Southeast Asia suggest that cancer was, when compared to other physical diseases, associated with the highest risk of suicide in older adults: in terms of magnitude, the risk of suicide linked to cancer was again on par with depression^10,11^

Relatively little is known about the impact of a new cancer diagnosis on risks of suicide among older adults. Three studies, one from the U.S.^12^, one from South Korea^13^, and one from Poland^14^ indicate, that old-age suicide risk is 1.6 to 2.6 times higher after cancer diagnosis. Given this limited evidence base from only three countries (with large differences in old-age suicide rates^15^), the high incidence of cancer among older adults^3^, the increase in the incidence of both cancer and suicide with age^5,16,17^, and increasing absolute numbers of both cancer and suicides in the foreseeable future due to population ageing^2,18^, further research into suicide risk after cancer diagnosis among older adults is needed. Particularly, it is important to identify high-risk groups and critical time windows to guide preventive efforts.

The aim of the current study was to analyse whether older adults with a new cancer diagnosis had a higher risk of suicide when compared to those who had not. Further, we assessed risks of suicide with regard to cancer site, cancer stage, and time since diagnosis. Newly available nationwide individual-level register data from Austria was used to examine this.

## METHODS

### Design, data sources and setting

We applied a cohort design to national data on all adults aged 65 years and older living in Austria, who were followed from 1^st^ of January 2014 until 31^st^ of December 2023. Individual-level, retrospective data from multiple registers (Central Population Register, Central Civil Status Register, Cancer Register) were obtained and linked via a pseudonymised identifier within the Austrian Micro Data Center of Statistics Austria. Information on cancer diagnoses were obtained from the Austrian National Cancer Registry (2014-2022), and data on causes of death came from the Central Civil Status Register (2014-2023), both employing the International Classification of Diseases, 10^th^ revision (ICD-10).

### Exposure

Exposure to a new cancer was identified as a new diagnosis with any cancer diagnosis (ICD-10: C00-C97) except for non-melanoma skin cancer (ICD-10: C44). The exact date when a new cancer was diagnosed was used as the index date. Individuals could be diagnosed with multiple new cancers during follow-up and, in these instances, the index date referred to the latest cancer diagnosis. When multiple cancers were diagnosed at the same day, we selected the diagnosis with the most advanced stage. For cancer site, we differentiated between the four most common types in Austria (prostate, breast, lung, and colorectal), a combined group of cancers with poor prognosis (pancreatic, liver, brain, oesophagus, stomach), as well as other cancers. Cancer stage was classified as stage 2 (local), stage 3 (regional), stage 4 (disseminated), or unknown.

### Outcome

The outcome was death by suicide and was identified based on ICD-10 codes of the underlying cause of death (ICD-10): X60-X84, Y87.0).

### Covariates

We adjusted for potential socio-demographic confounders, including age (in years), sex (men/women), level of education (coded according to International Standard Classification of Education (ISCED-1997) as low=1-2a/medium=3a-4c/high=5a-6), marital status (married/single/divorced/widowed), and area of living (urban, town, rural).

### Follow-up

Individuals newly diagnosed with cancer were followed from the date of diagnosis. For the rest of the older population without a new cancer diagnosis, an index date was allocated based on random draws from the index dates of cancer patients, so that the two groups had the same start of follow-up. Follow-up time was measured in 3-month intervals.^8^ All individuals were followed from the index date until they died by suicide, any other cause, or to the end of follow-up (five years), whichever came first.

### Statistical analysis

We calculated suicide incidence rates per 100,000 person-years for those newly diagnosed with cancer and those not diagnosed. Next, we estimated adjusted cumulative incidence functions for those with and without new cancer diagnosis using a non-parametric survival model (Aalen-Johansen estimator) with inverse probability treatment weighting (all standardised mean differences < 0.1) based on socio-demographic variables (age, sex, education, marital status, and area of living). Based on the adjusted cumulative incidence functions accounting for both competing risks (other causes of death) and confounding factors (differences between those with and without a new cancer diagnosis with regard to socio-demographics), risk ratios (relative risk) and risk differences (absolute risk) were calculated at multiple time points: 3 months, 1 year, 3 years, 5 years. All analyses were conducted in R (3.2.1)

## RESULTS

A total of 223,932 older adults were diagnosed with a new cancer between 2014 and 2022. The remaining older adults (n=1,951,202) were followed from a randomly assigned index date. Both groups were followed for up to five years. The mean follow-up time was 2.7 (SD=1.9) years among those diagnosed with a new cancer and 3.9 (SD=1.4) years among those without. During follow-up, 2,158 suicides occurred, of which 442 were among older adults newly diagnosed with cancer. The incidence rates were 74 and 23 per 100,000 person-years among those diagnosed and those not, respectively. Suicides were more common among older men and those aged 85 years and over (Table 1). 9% and 28% of suicides among cancer patients occurred within the first three months and the first year after diagnosis, respectively. The most common methods of suicide among older adults with a new cancer diagnosis were use of firearm (37%), hanging (36%), jumping from heights (9%), and poisoning (8%).

**Table 1:** Suicide deaths and incidence rates among older adults newly diagnosed with cancer and those not.

|  | Cancer diagnosis |  |  | No cancer diagnosis |  |  |
| --- | --- | --- | --- | --- | --- | --- |
|  | Suicides | Person-years | Incidence rate | Suicides | Person-years | Incidence rate |
| Total | 442 | 601,321 | 73.5 | 1716 | 7,633,490 | 22.5 |
| Sex |  |  |  |  |  |  |
| Male | 370 | 336,689 | 110.0 | 1,329 | 3,287,507 | 40.4 |
| Female | 72 | 264,633 | 27.2 | 387 | 4,345,983 | 8.9 |
| Age |  |  |  |  |  |  |
| 65-74 | 204 | 116,013 | 55.6 | 713 | 4,939,358 | 14.4 |
| 74-85 | 183 | 194,972 | 93.9 | 685 | 2,040,987 | 33.6 |
| ≥85 | 55 | 39,304 | 140.0 | 318 | 653,145 | 48.7 |

N=2,175,134, unweighted data. Incidence rates per 100,000 person-years.

After one year, risk for suicide was almost four times higher (RR=3.81, 95%-CI=3.19, 4.50) among those newly diagnosed with cancer versus those not when assessed in survival models that accounted for both competing risks and socio-demographic confounders (Table 2). Differences in relative risk within a year of diagnosis between males (RR=3.91, 95%-CI=3.15, 4.82) and females (RR=5.05, 95%-CI=3.12, 7.58) and between age groups (65-74 years: RR=5.20, 95%-CI=3.71, 7.05; 85+ years: RR=4.19, 95%-CI=2.57, 6.39) were limited and confidence intervals overlapped broadly. The highest risks were found among those with lung cancer (RR=4.20, 95%-CI=2.63, 5.83) and cancers with poor prognosis (RR=3.56, 95%-CI=2.22, 4.94). The lowest and not statistically significant excess risk was found for females with breast (2.22, 95%-CI=0.42, 4.16) and males with prostate cancer (1.23, 95%-CI=0.70, 1.78). With regard to cancer stage, the highest excess risk was found those with a disseminated (4.24, 95%-CI=3.07, 5.46) or unknown cancer stage (RR=4.10, 95%-CI=2.92, 5.33). Suicide risk was highest during the first three months (RR=7.57, 95%-CI=5.70, 10.07) and decreased thereafter, but remained elevated throughout follow-up (after 5 years: RR=1.71, 95%-CI=1.52, 1.90).

**Table 2:**
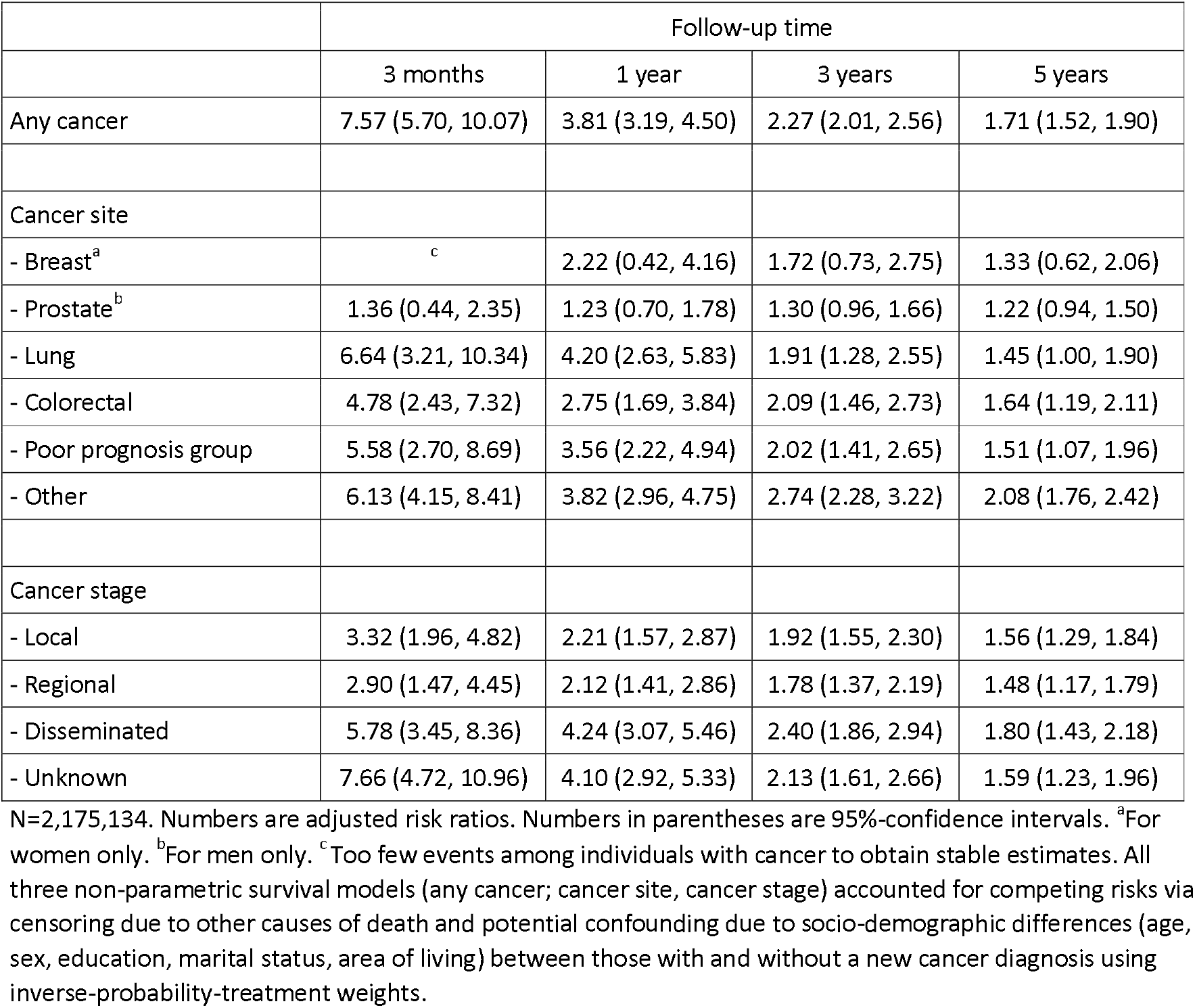
Adjusted risk ratios of a new cancer diagnosis for suicide death.

|  | Follow-up time |  |  |  |
| --- | --- | --- | --- | --- |
|  | 3 months | 1 year | 3 years | 5 years |
| Any cancer | 7.57 (5.70, 10.07) | 3.81 (3.19, 4.50) | 2.27 (2.01, 2.56) | 1.71 (1.52, 1.90) |
| Cancer site |  |  |  |  |
| - Breast<sup>a</sup> | <sup>c</sup> | 2.22 (0.42, 4.16) | 1.72 (0.73, 2.75) | 1.33 (0.62, 2.06) |
| - Prostate<sup>b</sup> | 1.36 (0.44, 2.35) | 1.23 (0.70, 1.78) | 1.30 (0.96, 1.66) | 1.22 (0.94, 1.50) |
| - Lung | 6.64 (3.21, 10.34) | 4.20 (2.63, 5.83) | 1.91 (1.28, 2.55) | 1.45 (1.00, 1.90) |
| - Colorectal | 4.78 (2.43, 7.32) | 2.75 (1.69, 3.84) | 2.09 (1.46, 2.73) | 1.64 (1.19, 2.11) |
| - Poor prognosis group | 5.58 (2.70, 8.69) | 3.56 (2.22, 4.94) | 2.02 (1.41, 2.65) | 1.51 (1.07, 1.96) |
| - Other | 6.13 (4.15, 8.41) | 3.82 (2.96, 4.75) | 2.74 (2.28, 3.22) | 2.08 (1.76, 2.42) |
| Cancer stage |  |  |  |  |
| - Local | 3.32 (1.96, 4.82) | 2.21 (1.57, 2.87) | 1.92 (1.55, 2.30) | 1.56 (1.29, 1.84) |
| - Regional | 2.90 (1.47, 4.45) | 2.12 (1.41, 2.86) | 1.78 (1.37, 2.19) | 1.48 (1.17, 1.79) |
| - Disseminated | 5.78 (3.45, 8.36) | 4.24 (3.07, 5.46) | 2.40 (1.86, 2.94) | 1.80 (1.43, 2.18) |
| - Unknown | 7.66 (4.72, 10.96) | 4.10 (2.92, 5.33) | 2.13 (1.61, 2.66) | 1.59 (1.23, 1.96) |

N=2,175,134. Numbers are adjusted risk ratios. Numbers in parentheses are 95%-confidence intervals. <sup>a</sup>For women only. <sup>b</sup>For men only. <sup>c</sup>Too few events among individuals with cancer to obtain stable estimates. All three non-parametric survival models (any cancer; cancer site, cancer stage) accounted for competing risks via censoring due to other causes of death and potential confounding due to socio-demographic differences (age, sex, education, marital status, area of living) between those with and without a new cancer diagnosis using inverse-probability-treatment weights.

The adjusted, absolute risk differences by cancer diagnosis and by sex and age sub-groups are shown in Figure 1. The strongest increase in suicide risk between those with and without new cancer diagnosis showed during the first year. After five years, the absolute risk of suicide death was 0.18% (95%-CI=0.16%, 0.20%) among those older adults who had received a new cancer diagnosis; implying that 0.18% had died by suicide with five years of the cancer diagnosis. Among those with no cancer, 0.11% (95%-CI=0.10%, 0.11%) died by suicide. The absolute risk of suicide among those newly diagnosed was higher among males than females, as well as among those aged 75 years and over versus those aged 65-74 years.

**Figure 1.**
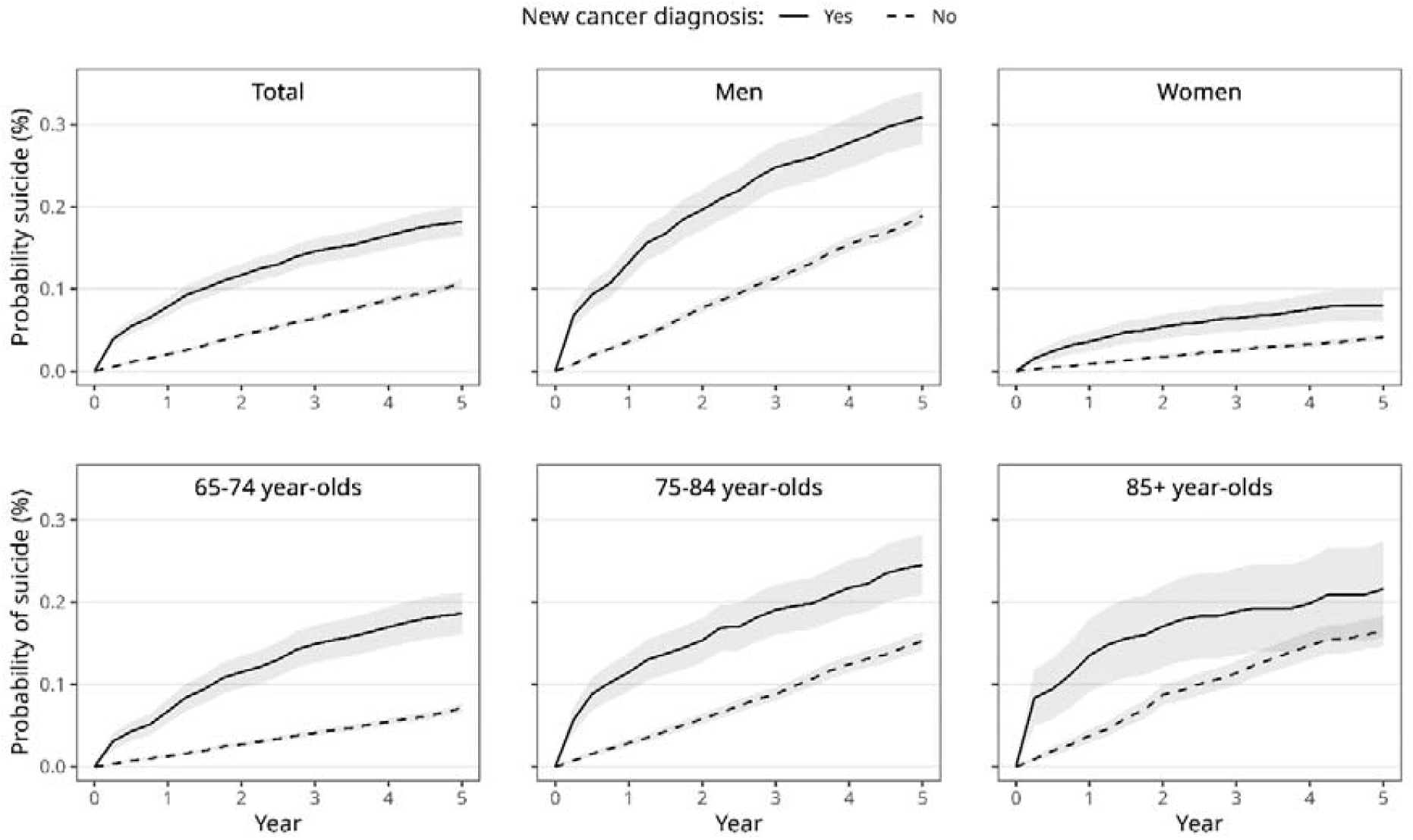
Adjusted probability of suicide death by whether a new cancer diagnosis was received or not N=2,175,134. Based on adjusted cumulative incidence functions and stratification estimated with non-parametric survival model stratified by sex and age which accounted for competing risks via censoring due to other causes of death and potential confounding due to socio-demographic differences between those with and without a new cancer diagnosis via inverse-probability-treatment weights.

## DISCUSSION

In this national, retrospective cohort study based on individual-level data, we found that older adults newly diagnosed with cancer had an excess risk of suicide when compared to those not diagnosed. The highest risks were found for lung cancer and those with a poor prognosis as well as those with disseminated cancer stages. The risk was particularly pronounced during the first three months after being diagnosed.

Our estimates were higher than pooled results from cancer-registry-based studies of all age groups^6,19^, including studies from Austria^20,21^. They were also higher than findings based on older adults in the US^12^, Poland^14^, and South Korea.^13^ A previous cohort study^13^ with a control group from the general population reported a two-fold higher risk among older adults within the first year of a new cancer diagnosis. However, the risk among Austrian older adults within one year after diagnosis was almost four-fold higher than in the comparison group. A potential explanations for these differences is hat studies based only on cancer-registry data, that is, when no individual-level data for the general population is available, could underestimate suicide risk due to the high prevalence of cancer^3,22^. Alternatively, we might also somewhat overestimate the relationship between cancer diagnosis and suicide as we adjusted for fewer potential confounders, at least when compared to studies based on more encompassing Nordic register data.^19^ We lacked specifically data on pre-cancer-diagnosis differences with regard to somatic comorbidities or psychiatric diagnoses. In the South Korean study^13^, cancer patients with a pre-diagnosed mental disorder had a higher risk (three-fold) to die by suicide compared to the majority of the population without such a diagnosis (two-fold). Similarly, a recent study^19^ reported a two-fold increased suicide risk among all cancer patients in Denmark compared to the general population after adjustment for pre-existing diagnosed psychiatric disorders. Finally, the relative suicide risk among older adults after cancer diagnosis in Austria could be truly higher than in other countries studied. We can only speculate, however, when it comes to the causes for any such deviation from other countries. The higher relative risk we found fits with the high absolute suicide rate among older adults in Austria compared to other European countries^2^, and the particularly high suicide rate among older men^15^, who constitute a vulnerable socio-demographic group also among cancer patients^23^. Compared to the recent Danish study^19^, for example, incidences rates of suicide among the oldest old (85+ years) in Austria were more than twice as high. Potential causal link factors as to why a cancer diagnosis may lead to suicide death likely include the initial psychological shock^24^, pain^25^ and other debilitating physical symptoms, loss of independence and physical functioning^26^, and inflammation^27^ due the disease and treatments, all of which may increase the subsequent risk for depression.

We found the risk of suicide peaked within the first three months after diagnosis and almost a third of suicide deaths of older adults with cancer occurred within the first year. This temporal effect has been reported also previously in all-age studies^5,6^, with estimates from studies ranging between a 12-fold risk within the first week^28^ to 4-5-fold within the first three months^8,28^. Again, our estimate of the risk during the first three months (7.5-fold) is somewhat higher than previous evidence. The higher risk directly after diagnosis is supported by the finding that cancer patients also have higher risk of depression in the first year after diagnosis.^29^ In line with results from studies among older individuals with cancer^13,14^, we also found a higher suicide risk for more advanced cancer stages as well as cancer sites with poorer prognosis. Although the design of our study does not allow to identify a causal effect, the temporal association combined with the consistent variation by prognosis supports a robust and specific link between specific cancer diagnoses and an increased suicide risk.

A clinical implication of our findings is that healthcare workers and relatives should be attentive of emotional distress, such as symptoms of anxiety, withdrawal, and depression, especially among older males already when being diagnosed and in the time shortly thereafter, especially if a cancer with a poor prognosis is diagnosed.^30,31^ As prediction of suicide is difficult given its rarity and the low sensitivity of common suicide risk factors^32^, screening for suicidal intent and psychological support should be considered for all cancer patients.

Strengths of this study include complete, nationwide registry data based on an individual-level linkage, thus, facilitating estimation of more precise risk estimates. Further, adjusting for relevant confounders reduced bias.

Limitations should be acknowledged as well. First, given that we followed the newly diagnosed for a maximum of five years and that we had no individual-level data on emigration, we might have underestimated the total suicide risk associated with cancer. However, studies^20,21^ with longer follow-up periods suggest only a limited underestimation, and emigration among older adults in the years 2019-2023 was limited (007E1%). Second, although the data quality of both the cancer registry^33^ and the cause of death register is considered to be reliable in Austria, we cannot exclude potential under-reporting of suicide death among older adults. Third, we did not have information on previous cancer diagnoses (before 2014) and focussed our analysis on the latest and most advanced new cancer diagnosis during follow-up. Hence, it remains unclear whether there is excess suicide risk in case of multiple (subsequent) cancer diagnoses. Fourth, data on diagnosed mental disorders was not available. This could be problematic, if the prevalence of mental disorders would vary substantively by cancer risk. A recent large individual participant meta-analysis^34^ concluded, however, that depression and anxiety disorder, the two most common mental disorders, were not associated with an increased cancer incidence, with the exception of a moderate increase with regard to smoking-related cancers. Fourth, we had no data on cancer-related treatment regime, hence we cannot differentiate between the effect of the cancer diagnosis itself and any (side-)effects of the subsequent treatment. However, since suicide risk is highest in the months immediately after diagnosis and diminishes subsequently, while treatments and their side-effects are often long-term^35^, the overwhelming initial shock of the cancer diagnosis appears more important for suicide deaths.^8^

In conclusion, we found that older adults who were newly diagnosed with cancer had a significantly higher risk of suicide than peers who had not been diagnosed. The risk of suicide was particularly high during the first three months after diagnosis as well as for lung cancer and cancers with poor prognosis. Early psychosocial support could help to address suicide risk among cancer patients.

## Author contributions

ES planned and conducted the analysis, and wrote the paper. AS, ELP, MAS, CW, CJ, TM, and AE critically reviewed the manuscript.

## Declarations of conflict of interest

None.

## Declarations of sources of funding

This work was supported by a grant from the Austrian Academy of Sciences (DATA_2023-08_SAOAA).

## Patient consent for publication

Not applicable.

## Ethics approval

The conduct of this study was approved by the Ethics Committee of the Medical University of Graz (1172/2024).

## Data

Access and linkage of register data (Central Population Register, Central Civil Status Register, Cancer Register, and Register-based Labour Market Statistics) was approved of and provided by the Austrian Micro Data Center (AMDC) of Statistics Austria following national legislation (Bundesstatistikgesetz §31, Forschungsorganisationsgesetz §38b). The AMDC is a research data infrastructure facility of Statistics Austria that enables research on micro data processed in compliance with data protection regulations. The data used for this research can be accessed by researchers at scientific institutions accredited with the AMDC against a fee.Forfurtherinformation,see https://www.statistik.at/en/services/tools/services/center-for-science/austrian-micro-data-center-amdc.

## Notes

### Competing Interest Statement

The authors have declared no competing interest.

